# Use of digital technology tools to characterize patient adherence to prescription-grade omega-3 polyunsaturated fatty acid therapy: Primary results of the DIAPAsOn prospective observational study

**DOI:** 10.1101/2022.02.09.22270713

**Authors:** Gregory P. Arutyunov, Alexander G. Arutyunov, Fail T. Ageev, Tatiana V. Fofanova

**Affiliations:** Department of Internal Medicine, Pirogov Russian National Research Medical University, Moscow.; Department of Outpatient Medical and Diagnostic Technologies, Myasnikov Institute of Clinical Cardiology, National Medical Research Center for Cardiology, Moscow.

**Keywords:** Omega-3-acid ethyl esters, myocardial infarction, hypertriglyceridemia, adherence, compliance, persistence, mhealth, ehealth, patient-reported outcomes

## Abstract

**Background, objectives and methods:** DIAPAsOn was a 6-month prospective observational multicenter study in the Russian Federation that examined adherence to a preparation of highly purified omega-3 polyunsaturated fatty acids (OMACOR) in adult patients with a history of recent myocardial infarction or endogenous hypertriglyceridemia. A full description of the study’s aims and methods has appeared in *JMIR Res Protoc.* A feature of DIAPAsOn was the use of a bespoke electronic patient engagement and data collection system.

**Results:** The net average reduction from baseline in both total and low-density lipoprotein cholesterol was approximately 1 mmol/L and the net average increment in high-density lipoprotein cholesterol was 0.2±0.53 mmol/L. Mean triglyceride levels declined by ∼1.3 mmol/, from an initial level of 2.99±1.29 mmol/L to 1.67± 0.67 mmol/L. The percentage of patients with triglyceride <1.7 mmol/L rose from 13.1% at baseline to 54% at study-end.

Digital reporting of adherence was registered by 8.3% of patients (n=180) and average scores indicted poor adherence. However, clinic-based enquiry suggested high levels of adherence.

Data on health-related quality of life accrued from digitally-engaged patients identified improvements among patients reporting high adherence to study treatment, but patient numbers were small.

**Conclusions:** The lipid and lipoprotein findings indicate that OMACOR had nominally favorable effects on the blood lipid profile. Less than 10% of patients enrolled in DIAPAsOn used the bespoke digital platform piloted in the study and the level of self-reported adherence to medication by those patients was also low. Reasons for this low uptake and adherence are unclear. Better adherence was recorded by clinical report.

## Introduction

Non-high-density lipoprotein cholesterol blood lipids are a source of residual cardiovascular risk in patients whose low-density lipoprotein cholesterol (LDL-C) levels are well controlled by medication, primarily statins [1–9].

Optimal risk reduction in cardiovascular disease, as in other forms of major non-communicable diseases, depends substantially on patients continuing to take their medications for extended periods of time. This can be a particular challenge in conditions such as hyperlipidemia, where the connection between symptomless elevations of blood lipid levels and major cardiovascular events can seem abstract or remote [10, 11].

Recent comparative research in Russia and Norway has disclosed poor attainment of cholesterol targets in both countries, despite a notably higher prescription rate of these drugs in Norway [12]. Suboptimal patient adherence to prescribed treatments is likely to be a contributor to such findings, which illustrates that the challenges of promoting and sustaining adherence to therapy are not confined to any one country. It is nevertheless clear from the results of the CEPHEUS II study that failure to reach targets for lipid-based risk reduction is widespread in Russia [13]. Patient-related factors associated with non-attainment of targets identified in that study included considering it acceptable to miss prescribed doses more than once per week. Poor adherence to medication for hypertension has likewise been documented in the Izhevsk Family Study II [14].

Those findings exemplify observations that the rates of both discontinuation and non-adherence to therapy are uniformly high in clinical trials of lipid-lowering drugs and even higher in unselected populations, with adherence deteriorating in proportion to the duration of follow-up [15]. Analysis of a large Swiss healthcare claims database (N=4349) revealed that overall adherence to drug therapy for secondary cardiovascular prevention after myocardial infarction (MI) was only moderate, but that patients with high adherence to lipid-lowering therapy had a significantly reduced risk for all-cause mortality and major cardiovascular events, illustrating the potential for improvement of longer-term outcomes [16].

OMACOR (hereafter OM3EE; license holder Abbott Laboratories GmbH) is a preparation of highly purified long-chain omega-3 polyunsaturated fatty acids (n-3 PUFAs) (eicosapentanoic acid/docosohexanoic acid in a 1.2:1 ratio and 90% purity) available as a prescription-only medication and widely approved for use, at a daily dose of 1 g, for the secondary prevention of major cardiovascular events in patients who have survived an MI, or at doses of 2–4 g/day for the regulation of triglycerides (TG). Prescription-only n-3 PUFAs (PnP) such as OM3EE are qualitatively distinct from dietary n-3 PUFA supplements and have been evaluated in a range of clinical trials [17, 18].

The emergence of widely available digital and internet technologies with the potential to provide immediate bidirectional communication between healthcare professionals (doctors, nurses and pharmacists) and patients may be an important new resource for promoting long-term adherence to therapies [19]. The DIAPAsOn study was devised to explore patient adherence to OM3EE therapy through the medium of digital technology tools.

## Methods

### Overview

A comprehensive description of the methodology of the DIAPAsOn study has been published, including baseline demographic data [20].

Briefly, DIAPAsOn was a prospective observational study conducted at >100 centers in the Russian Federation and devised to examine adherence to a prescription of OM3EE as either a secondary preventive medical therapy (at a dose of 1 g/day) for patients with a history of recent MI or for blood lipid regulation (at doses of 2–4 g/day) in patients with endogenous hypertriglyceridemia insufficiently responsive to dietary modification or drug therapy.

Participants were required to be adults (≥18 years) with a history of MI for whom OM3EE was prescribed as part of a secondary prevention strategy or to have Fredrickson endogenous type IIb or III hypertriglyceridemia not satisfactorily controlled by statin therapy or Fredrickson endogenous type IV hypertriglyceridemia not sufficiently controlled by a lipid-moderating diet. In addition, patients should have been taking OM3EE for not >2 weeks prior to enrolment.

DIAPAsOn is registered at www.ClinicalTrials.gov (NCT03415152).

### Schedule of visits and data collection

The DIAPAsOn study had a scheduled duration of 6 months. Clinic visits were scheduled at the start of the study (Visit 1) and then at approximately 3 months (Visit 2) and at the end of the study (Visit 3). At each of the three scheduled clinic visits, patients were questioned about compliance with OM3EE therapy using the Questionnaire of Treatment Compliance [21]. This instrument, which has been used in Russia to investigate compliance with other cardiovascular medications, produces a numerical indication of compliance, as follows: 12–15 points, very high; 8–11 points, high; 4–7, moderate; and 0–3, low.

Blood lipid profile was determined at each visit, along with blood pressure and heart rate data, adverse events and hospitalization were recorded. Patients also received inter-visit phone calls focused on adherence to therapy and safety.

A central aspect of DIAPAsOn was the use of remote digital technology that allowed patients to submit data and report on matters such as health-related quality of life (HRQoL) and product usability (very good, good, moderate, poor).

The electronic patient engagement and data collection system used in DIAPAsOn was developed in collaboration with the medical online platform ROSMED.INFO (Moscow, Russian Federation; www.rosmed.info), which has wide-ranging experience in the development and operation of mobile health applications in the Russian Federation. A fuller description of the system used in DIAPAsOn featured in the separate publication of methodology [20].

All aspects of the DIAPAsOn study, including the associated mobile health app, conformed to relevant extant national and international legal and ethical regulations and requirements for the conduct of clinical research in human subjects, including the provisions of the Declaration of Helsinki, including patients’ right to decline further participation in DIAPAsOn at any time and for any reason, whether stated or not, without prejudice to their subsequent treatment. Ethical oversight of the DIAPAsOn study was exercised by the independent Interuniversity Ethics Committee (Moscow, Russian Federation). A list of center investigators appears in *Appendix 1*.

### Statistical methods

Methods were predominantly descriptive, conducted in accordance with the pre-approved statistical analysis plan, and used the statistical programming language R (version 3.4.3; https://mran.microsoft.com/releases/3.4.3).

The primary endpoint—adherence to therapy with OM3EE in post-MI patients or patients with hypertriglyceridemia—was assessed in an analysis population, defined as those patients for whom data were obtained at least at Visits 1 and 2.

Analysis of the primary endpoint included determination at the end of the study (Visit 3) of the mean adherence rate, which was defined as the number of days for which the patient took the full prescribed dose of OM3EE during the specified period divided by the total number of days in that period. The mean score on the National Questionnaire of Treatment Compliance was calculated at the same time.

Comparison of individual patient data between visits was based on either Student’s t-test (for dependent variables) or McNemar’s test (for qualitative data).

## Results

A total of 3000 patients were initially included in the program but 428 (14.3%) were excluded because Visit 1 data were incomplete. Valid and complete data from Visit 1 were available for 2572 patients (85.7%), who constituted the safety population. After exclusion of 405 patients lost before Visit 3, an analysis population of 2167 patients was derived, representing 72.2% of total enrolled patients (Figure 1), of whom 898 (41.4%) were taking OM3EE for secondary prevention after MI and 1269 (58.6%) were taking OM3EE for hypertriglyceridemia.

**Figure 1.**
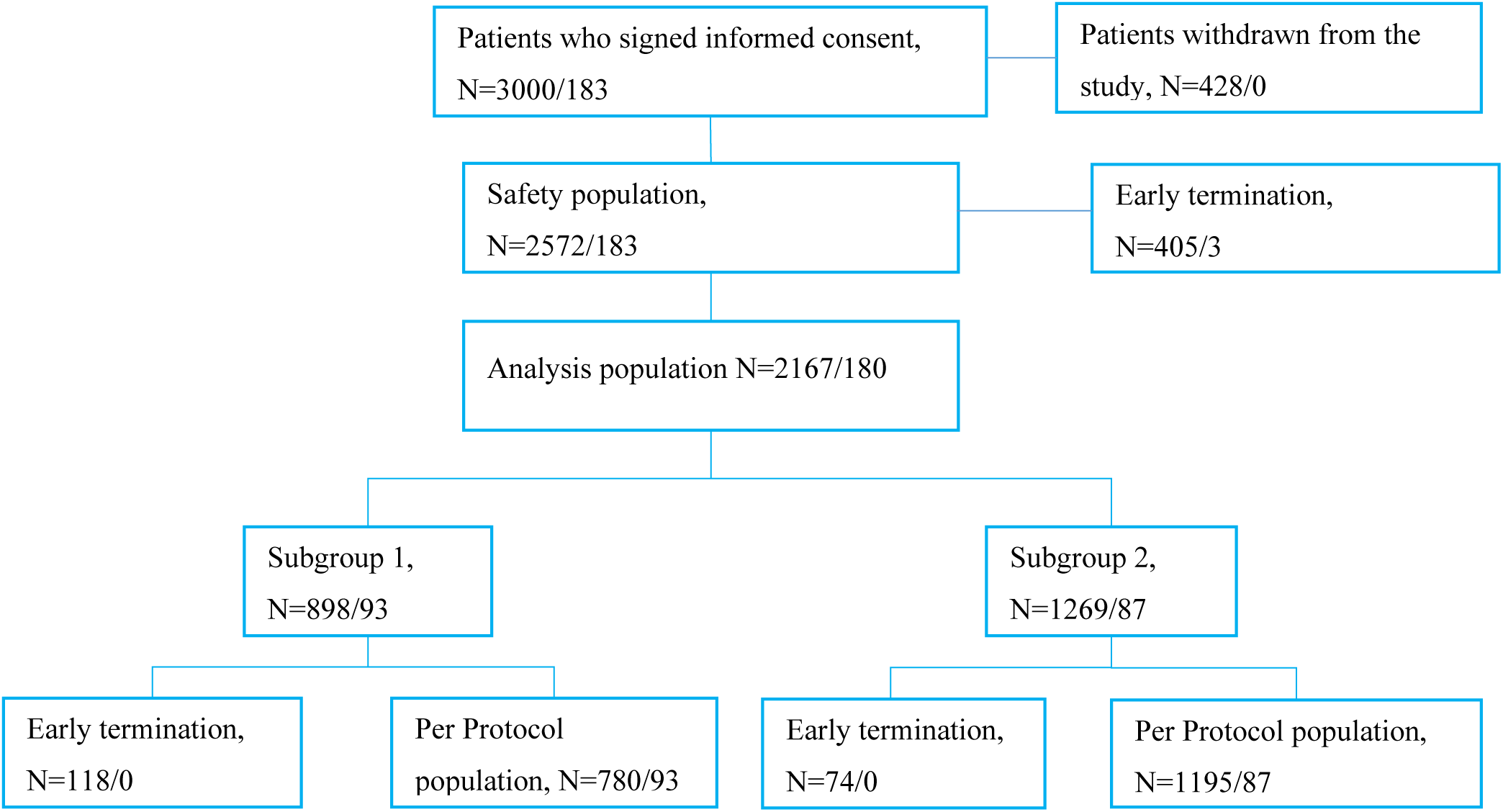
Derivation of patient subsets in the DIAPAsOn study.

DIAPAsOn was completed per protocol by 1975 patients (secondary prevention, n=780; hypertriglyceridemia, n=1195). This was an almost wholly Caucasian population (97.74%) with a near-equal sex distribution (men, 52.8%; women, 47.2%), an average age of ≍60 years and an average body mass index (BMI) of 30 kg/m^2^. There were more men than women in the secondary prevention subgroup (n=608; 67.7%), whereas women outnumbered men in the hypertriglyceridemia subgroup (n=732; 57.68%). Investigator-assessed ‘clinically significant’ abnormalities of systolic and diastolic blood pressure (BP) were recorded in 21.4% and 12.8% of patients, respectively.

As illustrated in Figure 1, 180 of the 2167 patients who comprised the analysis population (8.3%) submitted data via the mobile health platform, of whom 93 were enrolled in DIAPAsOn on the basis of previous MI and 87 on a diagnosis of qualifying hyperlipidemia. From start to finish, three of the initial grouping of 183 patients who used the mobile health platform (1.6%) were withdrawn from the study or discontinued it, compared with 1025 of 2817 (36.4%) of those who did not submit data via the mobile platform. After establishment of the analysis population, early termination rates were 0% and 9.7%, respectively (**Figure 1**).

### Compliance with OM3EE

The mean duration of OM3EE administration was 166.5±70.6 days (median, 199 days; range, 14–268 days).

Among the 2167 patients whose compliance was monitored at clinical visits but not self-reported via the mobile app, the mean score on the National Questionnaire of Treatment Compliance at Visit 3 was 13±3 points, signifying high overall compliance with therapy. Mean scores >12 were also recorded at Visit 1 (12.5±3.1) and Visit 2 (13±2.9). Relative to the mean score at Visit 1, the mean scores at both Visits 2 and 3 were statistically significantly larger (p<0.001) (Table 1). The distribution of adherence categories for the total study population and for the two subpopulations of DIAPAsOn is shown in Figure 2. Overall, high or very high compliance was recorded for >90% of respondents in both subseries.

**Figure 2.**
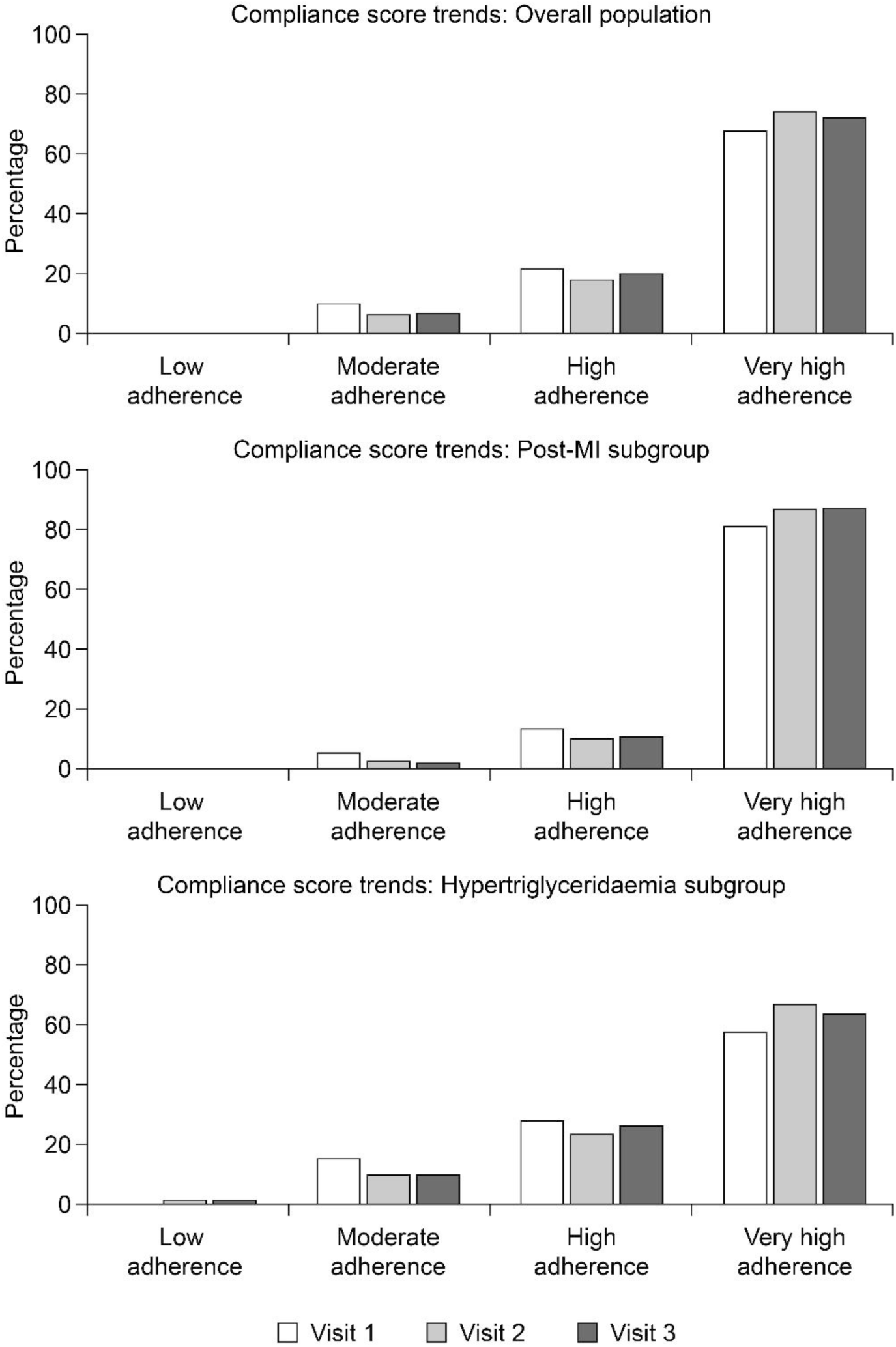
Distribution of adherence categories for the total study population and for the two subpopulations (post-MI and hypertriglyceridemia) of the DIAPAsOn analysis population, based on responses to the National Questionnaire of Treatment Compliance.

**Table 1.**
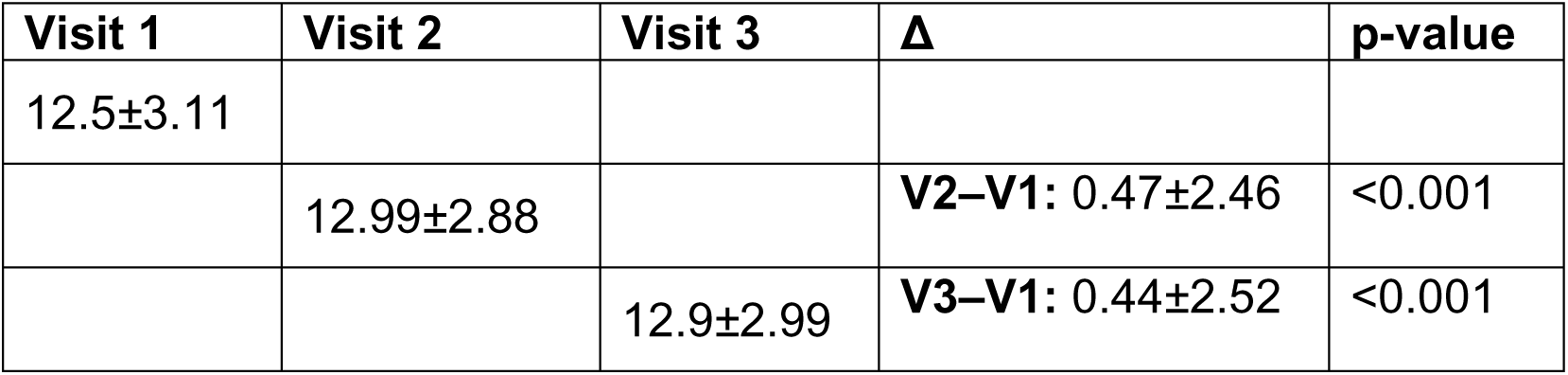
Trends in mean score on the National Questionnaire of Treatment Compliance between baseline (Visit 1) and Visit 2 (3 months of study participation) or Visit 3 (study completion, after 6 months).

| Visit 1 | Visit 2 | Visit 3 | $\Delta$ | p-value |
| --- | --- | --- | --- | --- |
| 12.5±3.11 | | | | |
| | 12.99±2.88 | | **V2–V1:** 0.47±2.46 | <0.001 |
| | | 12.9±2.99 | **V3–V1:** 0.44±2.52 | <0.001 |

A total of 69 patients (2.68%) discontinued OM3EE during the study. Numerically, the largest groups to do so were patients who cited inconvenience of use (n=11) and those who reported absence of stock at pharmacies (n=6). A total of 50 patients discontinued for a plurality of reasons, including cost of medication, reluctance to commit to long-term medication discontinued by another physician, normalization of blood lipid values and change of residence.

Inconvenience of use was more often recorded in patients enrolled in DIAPAsOn for hypertriglyceridemia than for secondary prevention (19% vs 5.9% of total cases).

Among the 180 patients who registered data via the DIAPAsOn mobile platform, adherence to therapy, expressed as the ratio of days when the full prescribed dose of OM3EE was taken to the total number of days in the treatment period, averaged 0.37±0.38 over the entire program, corresponding to a low level of adherence. Mean adherence between Visits 1 and 2 was 0.48±0.4, while that between Visits 2 and 3 was 0.24±0.4 (p<0.001). Between Visits 1 and 2, 50.0% of patients had low adherence (<0.5), 15.56% had moderate adherence (0.5–0.7) and 34.44% had high adherence (≥0.8). Between Visits 2 and 3, the proportion of patients with low adherence increased to 75.0%, while the proportion with high adherence decreased to 20.56%. When the data were stratified by adherence level there was an essentially binary split, with most patients reporting either low adherence (74.44%) or high adherence (25%).

In the subgroup of 93 patients taking OM3EE for secondary prevention after MI and self-reporting adherence via the study app, mean adherence between Visits 1 and 3 was 0.47±0.39, with 66.67% of patients recording low adherence and 32.26% high adherence. Mean adherence in that subgroup at Visit 2 reached 0.6±0.38; at Visit 3 it had decreased to 0.33±0.45 (p<0.001 for trend). At Visit 2, 32.26% of patients had low adherence (<0.5) and 45.16% had high adherence (≥0.8). By Visit 3, the proportion of patients with low adherence had increased to 67.74%, while the proportion with high adherence had declined to 30.11%.

Among the 87 app-using patients taking OM3EE for hypertriglyceridemia, mean adherence between Visits 1 and 3 was 0.25±0.33, with most patients (82.76%) self-reporting low adherence, and 17.24% recording high adherence. In this subgroup, 68.97% of patients had low adherence (<0.5) at Visit 2 and 22.99% had high adherence (≥0.8). By Visit 3, those percentages had changed to 82.76% and 10.34%, respectively.

Cross-referencing of the results on the National Questionnaire of Treatment Compliance administered at clinic visits with self-reported adherence based on the ratio of administered and prescribed dose established that, among patients identified by their response to the National Questionnaire as having ‘very high’, ‘high’ or ‘moderate’ adherence to therapy, app-reported mean adherences for the time period between Visits 1 and 2 were 50.04%, 52.85% and 24.58%, respectively, while between Visits 2 and 3 adherence was 28.67% for those assessed as having ‘very high’ adherence by questionnaire, 19.11% for those in the ‘high’ category and 5.57% for those in the ‘moderate’ category.

In the analysis population as a whole, 64.5% of patients rated the usability of OM3EE after 1 month of treatment as ‘very good’. A further 29.9% and 5.6%, respectively rated usability as ‘good’ or ‘moderate’. No patient rated usability as ‘poor’. All the patients prescribed OM3EE for secondary prevention post-MI rated the usability as ‘very good’ (70.7%) or ‘good’ (29.3%), while, among patients treated for hypertriglyceridemia, the usability of OM3EE was rated as ‘very good’ by 57.1% of patients, ‘good’ by 30.6% and ‘moderate’ by 12.2%.

### Lipid indices

At baseline, investigator-classified ‘clinically significant’ deviations from normal for total cholesterol (TC), LDL-C, high-density lipoprotein cholesterol (HDL-C) and TG were recorded in 46.1%, 40.93%, 14.4% and 64.97% of patients, respectively. Mean values of TC, TG, LDL-C, HDL-C and non-HDL-C (n-HDL-C) were 5.55±1.39, 2.99±1.29, 3.5±1.25, 1.27±0.46 and 4.29±1.47 mmol/L, respectively.

In-study trends in mean blood lipid levels are depicted in Figure 3 for the overall DIAPAsOn cohort and for the two subpopulations differentiated by indication.

**Figure 3.**
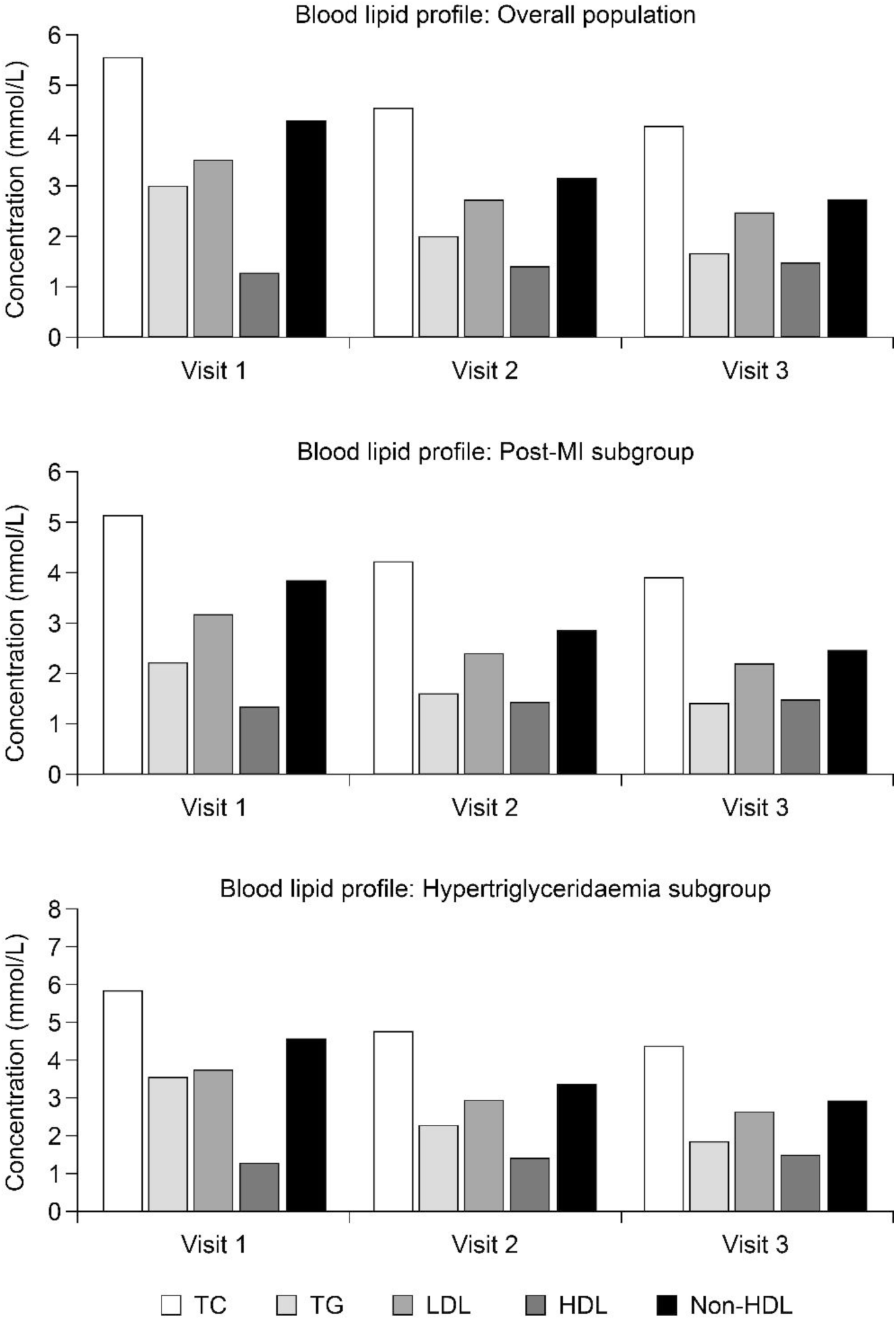
In-study trends in blood lipids in the total study population and for the two subpopulations (post-MI and hypertriglyceridemia) of the DIAPAsOn analysis population.

Analysis of lipid profile stratified by baseline TG status revealed that increasing TG levels were associated with trends in TC that were potentially deleterious to cardiovascular health (Table 2).

**Table 2.** Baseline lipid profile of the analysis population, stratified by TG status, identified progressively more atherogenic patterns of total and lipoprotein cholesterol as TG levels increased.

| **TG**<br>**(mmol/l)** |  | **TC (mmol/l)** | **TG (mmol/l)** | **LDL-C**<br>**(mmol/l)** | **HDL-C**<br>**(mmol/l)** | **n-HDL-C**<br>**(mmol/l)** |
| --- | --- | --- | --- | --- | --- | --- |
| <1.7 | N | 282 | 282 | 282 | 282 | 280 |
|  | Mean | 4.67 | 1.21 | 2.93 | 1.33 | 3.37 |
|  | SD | 1.33 | 0.31 | 1.14 | 0.5 | 1.4 |
|  | Median | 4.5 | 1.2 | 2.7 | 1.2 | 3.25 |
| 1.7–2.3 | N | 431 | 431 | 430 | 431 | 431 |
|  | Mean | 5.31 | 2.04 | 3.39 | 1.31 | 4 |
|  | SD | 1.24 | 0.19 | 1.3 | 0.51 | 1.4 |
|  | Median | 5.5 | 2.01 | 3.1 | 1.2 | 4.01 |
| >2.3 | N | 1438 | 1438 | 1438 | 1438 | 1437 |
|  | Mean | 5.79 | 3.63 | 3.65 | 1.25 | 4.55 |
|  | SD | 1.37 | 1.08 | 1.22 | 0.44 | 1.42 |
|  | Median | 5.9 | 3.4 | 3.5 | 1.1 | 4.6 |

Lipid profile parameters were recorded at baseline at Visit 1, as well as after 3 months (Visit 2) and 6 months (Visit 3) of follow-up. Mean TC at Visit 1 was 5.55±1.39 mmol/L. This had decreased to 4.54±1.04 mmol/L (p<0.001) by Visit 2, and at Visit 3 had been further reduced, to 4.17±1.04 mmol/L (p<0.001). Across the period of observation, the net average change in mean TC was thus –1.32±1.28 mmol/L.

At Visit 2, mean LDL-C was 2.71±0.94 mmol/L, an average reduction from Visit 1 of 0.77±0.92 mmol/L (p<0.001). Further reduction was observed at Visit 3, when the mean LDL-C level was 2.46±0.76 mmol/L (p<0.001 vs Visit 1). The average net decrement in LDL-C was thus 1.02±1.02 mmol/L.

Mean HDL-C levels at Visits 1, 2 and 3 were, respectively, 1.27±0.46, 1.41±0.42 (p<0.001 vs Visit 1) and 1.44±0.42 mmol/L (p<0.001 vs Visit 1), with an average increment of 0.2±0.53 mmol/L over the period of observation.

n-HDL-C declined by an average of 1.6±1.54 mmol/L during the period of observation, falling from 4.27±1.47 mmol/L at Visit 1 to 3.14±1.12 mmol/L at Visit 2 (p<0.001 vs Visit 1) and to 2.71±1.0 mmol/L at Visit 3 (p<0.001 vs Visit 1).

Mean TG was 3.0±1.3 mmol/L at Visit 1, 2.0±0.9 mmol/L at Visit 2 and 1.7±0.7 mmol/L at Visit 3 (p<0.001 for both vs Visit 1). The overall average reduction in mean TG was thus 1.32±1.15 mmol/L across the observation period.

Trends in overall TG levels during DIAPAsOn are displayed in more detail in Table 3, with patients assigned to one of three baseline distribution categories. Statistically significant changes in that distribution towards lower levels of TG were apparent at both Visits 2 and 3, with the percentage of patients recorded as having TG <1.7 mmol/L increasing from 13.1% at baseline to 54% at the conclusion of the study period, while the percentage recorded as having TG >2.3 mmol/L fell from 66.9% to 10.4%.

**Table 3.** Trends in overall TG levels during DIAPAsOn, stratified by baseline TG category.

| Visit | Parameter | Baseline TG (mmol/l) |  |  |
| --- | --- | --- | --- | --- |
|  |  | <1.7 | 1.7–2.3 | >2.3 |
| 1 | N | 2151 |  |  |
|  | n/% | 282/13.1 | 431/20 | 1438/66.9 |
| 2 | N | 2037 |  |  |
|  | n/% | 787/38.6 | 670/32.9 | 580/28.5 |
|  | p-value Visit 2–Visit 1 | <0.001 | <0.001 | <0.001 |
| 3 | N | 1905 |  |  |
|  | n/% | 1028/54 | 679/35.6 | 198/10.4 |
|  | p-value Visit 3–Visit 1 | <0.001 | <0.001 | <0.001 |

The average differences in TC, LDL and n-HDL-C between baseline and Visit 3 were a function of baseline TG. Thus, the reductions in patients with initial TG >2.3 mmol/L were –1.47, –1.1 and –1.7 mmol/L, respectively, while in patients with initial TG 1.7–2.3 mmol/L, the average intra–study changes from baseline to Visit 3 were –1.16, – 1.99 and –1.32 mmol/L. In patients with initial TG <1.7 mmol/L the average changes in TC, LDL and n-HDL-C were –0.83, –0.63 and –0.94 mmol/L, respectively. Significance was nevertheless apparent (p<0.001) for each parameter in each of those subgroups. The mean change in HDL during observation versus baseline was +0.24 mmol/L in patients with TG >2.3 mmol/L (p<0.001), +0.16 mmol/L in patients with TG 1.7–2.3 mmol/L (p<0.001) and +0.09 mmol/L in patients with TG <1.7 mmol/L (p=0.002). A statistically significant decrease in TG at Visit 3 versus baseline was observed only in subgroups of patients with baseline TG >2.3 or 1.7–2.3 mmol/L (–1.81 and –0.55 mmol/L, respectively; p<0.001 for both subgroups).

Sub-analysis of the patients being treated for hypertriglyceridemia stratified according to the concomitant use/non-use of statins and/or fibrates identified no substantial or significant inter-group differences in baseline levels of blood lipid components. Subsequent in-study trends in blood lipid fractions in both these subgroups are summarized in Table 4 and indicate significant longitudinal trends in both subgroups (p<0.001 for all indices in both comparisons) and slightly more pronounced responses in those who were taking additional lipid-regulating drugs in combination with OM3EE.

**Table 4.** Trends in lipid and lipoprotein fractions in patients enrolled in the analysis population of DIAPAsOn for hypertriglyceridemia and receiving or not receiving concomitant statins and/or fibrates.

|  | **TC**<br>**(mmol/l)** | **TG (mmol/l)** | **LDL-C**<br>**(mmol/l)** | **HDL-C**<br>**(mmol/l)** | **n-HDL-C**<br>**(mmol/l)** |
| --- | --- | --- | --- | --- | --- |
|  | **Statins or fibrates: YES** |  |  |  |  |
| V3–V1<br>mean±SD | –1.5±1.3 | –1.7±1.2 | –1.2±1.1 | +0.2±0.5 | –1.8±1.5 |
| V3–V1<br>median | –1.44 | –1.7 | –1.0 | +0.2 | –1.9 |
| P-value (V3<br>vs. V1)* | 0.001 | 0.001 | 0.001 | 0.001 | 0.001 |
|  | **Statins or fibrates: NO** |  |  |  |  |
| V3–V1<br>mean±SD | –1.4±1.3 | –1.5±1.1 | –0.84±0.9 | 0.3±0.6 | –1.7±1.6 |
| V3–V1<br>median | –1 | –1.32 | –0.7 | +0.2 | –1.3 |
| P-value (V3<br>vs. V1)* | 0.001 | 0.001 | 0.001 | 0.001 | 0.001 |

Formal tests for inter-subgroup differences depending on the use/non-use of statins or fibrates were not conducted.

Changes between baseline and Visit 3 stratified by self-reported adherence registered via the study mobile app (n=180) identified no correlations or associations between the level of adherence and absolute changes in levels of lipids or lipoproteins. The results of investigations into the relationship between rates of adherence and patient demographic factors for the whole analysis population are summarized in the *Supplementary Tables*.

### HRQoL outcomes

HRQoL data accrued from patients who contributed to the digital data collection element of DIAPAsOn are summarized in Table 5. Data for the ‘General Health’ domain were excluded due to a technical error during data transfer. Statistically significant increases in the scores for all domains except pain were recorded during the observation period. Further analysis, stratified by self-reported adherence to therapy (low, moderate or high), indicated that these improvements in HRQoL were restricted to patients with high compliance (data not shown; n=21–35).

**Table 5.** HRQoL data accrued from patients who contributed to the digital data-collection element of DIAPAsOn. Differences (Visit 2-Visit 1) and (Visit 3-Visit 1) were paired and therefore estimated only for those patients who were scored on both relevant visits. Data for the ‘General Health’ domain were excluded due to a technical error during data transfer.

| Visit | Parameter | PF | RP | RE | E/F | EW | SF | P |
| --- | --- | --- | --- | --- | --- | --- | --- | --- |
| 1 | N | 82 | 82 | 82 | 82 | 82 | 82 | 82 |
|  | Mean | 22.36 | 39.02 | 53.25 | 43.82 | 52.9 | 58.54 | 74.45 |
|  | SD | 16.18 | 44.11 | 41.2 | 21.62 | 16.1 | 26.34 | 31.16 |
| 2 | N | 51 | 50 | 50 | 49 | 49 | 50 | 49 |
|  | Mean | 34.56 | 87.5 | 93.33 | 72.28 | 76.01 | 89.25 | 94.23 |
|  | SD | 12.34 | 29.99 | 20.2 | 14.09 | 15.7 | 17.31 | 13.19 |
| 2–1 | N | 35 | 35 | 35 | 35 | 35 | 35 | 35 |
|  | Mean | 7.63 | 32.86 | 25.71 | 20.81 | 22.66 | 26.43 | 8.43 |
|  | SD | 13.4 | 48.42 | 32.42 | 14.12 | 14.69 | 25.68 | 14.12 |
|  | p-value* | 0.002 | <0.001 | <0.001 | <0.001 | <0.001 | <0.001 | 0.001 |
| 3 | N | 22 | 22 | 22 | 22 | 22 | 22 | 22 |
|  | Mean | 40.1 | 98.86 | 100 | 80.23 | 86.73 | 98.3 | 98.52 |
|  | SD | 6.42 | 5.33 | 0 | 5.87 | 7.94 | 8 | 6.93 |
| 3–1 | N | 22 | 22 | 22 | 22 | 22 | 22 | 22 |
|  | Mean | 6.69 | 29.55 | 18.18 | 18.18 | 25 | 16.48 | -0.11 |
|  | SD | 10.08 | 43.39 | 30.39 | 13.59 | 14.65 | 25.41 | 0.53 |
|  | p-value* | 0.005 | 0.004 | 0.011 | <0.001 | <0.001 | 0.006 | 0.329 |

### Safety and adverse events data

A safety population was identified that included all patients who had completed at least Visit 1 (n=2572).

A total of four adverse drug reactions (ADRs) were recorded in three patients (0.12%). Two patients had one ADR and one patient had two. No serious ADRs were recorded during DIAPAsOn.

Four deaths were recorded during the study, including one from cardiovascular disease. None of those deaths were causally related to use of OM3EE.

There were 20 instances of hospitalization due to cardiovascular diseases, none of which were attributed to use of OM3EE. Thirteen of these events affected participants who were being treated for hypertriglyceridemia, all of whom were also being medicated with statins and/or fibrates.

OM3EE therapy was discontinued by 69 patients. Specified reasons for doing so comprised inconvenience of use (n=11), lack of availability in pharmacies (n=6) and lack of effect (n=2). Reasons for the remaining 50 discontinuations were recorded as ‘other’.

## Discussion

Considered overall, the data from DIAPAsOn suggest that the introduction of OM3EE had favorable effects on the blood lipid profile of our patients, consistent with experience in previous controlled trials. As illustrated in Table 3 the percentage of patients recorded as having TG <1.7 mmol/L quadrupled in response to OM3EE (from 13.1% at baseline to 54% at the conclusion of the study); conversely the percentage of patients recorded as having TG >2.3 mmol/L fell to 10.4% from 66.9% at baseline. These changes were accompanied by alterations in other lipoprotein fractions compatible with an overall shift to a less atherogenic lipid profile, including reductions in non-HDL-C which declined by an average 1.6±1.54 mmol/l. This pattern of response to of OM3EE was substantially independent of the use or non-use of statins by patients treated for hypertriglyceridemia (Table 4).

By implication, the levels of treatment adherence in both the digital and non-digital contingents of DIAPAsOn were sufficient to achieve those effects, notwithstanding discrepancies in the level of adherence reported by the two groups of patients.

Among the analysis population in whom adherence was determined by in-clinic questioning adherence was consistently substantially higher in the post-MI subset than the hypertriglyceridemia subset (Δ 10–20%). As survivors of a major cardiac episode, those patients may be more motivated to continue with their medications than patients with an invisible and asymptomatic condition. This indication of a motivation element based on patients’ clinical history is consistent with the findings of Gaisenok et al. in the PROFILE registry [22], but not wholly sustained by the findings of the ISLAND study [23]. Within the post-MI group adherence fell significantly at age >75 years (75.31% vs >87.5% for all younger age deciles; p=0.007, chi-square test).

Frequency of dosing may also be an influence on the discrepancy in adherence between our two patient subgroups. The prescribed dose of OM3EE for secondary prevention post-MI is 1 g/day, whereas that for hypertriglyceridemia is 2–4 g/day. The up to fourfold difference in acquisition costs, plus the necessity of a twice-daily (vs once-daily) dosing regimen, may have contributed to the lower adherence recorded in the hypertriglyceridemia subgroup of DIAPAsOn, and perhaps to patients’ views on the usability of OM3EE. However, the detailed patient interviews required to elucidate this matter were not part of the study program.

‘Very high’ adherence was reported significantly more often by men than women, especially in the hypertriglyceridemia subset (67.71% vs 60.38%; p=0.007, chi-square test). Among patients with hypertriglyceridemia, ‘very high’ adherence was also significantly more likely in those who were recorded as not working (69.23% vs 59.66%; p<0.001, chi-square test), suggesting perhaps an effect of competing priorities. Adherence was much higher among early school leavers than in any other category of education but, especially in the hypertriglyceridemia subset, this finding was based on small numbers (n=3).

A further potentially strong influence on patient adherence and persistence is the personal beliefs of health professionals and general population attitudes. ‘Therapeutic inertia’ among physicians has been reported to contribute to poor management of cardiovascular risk factors in Russia and the USA [24, 25]. The fact that 31% of the post-MI subgroup of DIAPAsOn were classified as current smokers may be indicative of attitudinal factors that operate to the general detriment of adherence.

A central purpose of DIAPAsOn was to examine how the use of digital technologies might promote adherence to OM3EE therapy. This aspect of the study provides inconclusive and somewhat perplexing insights. The online facilities developed for DIAPAsOn were used by 180 of the 2167 patients (8.3%) of the analysis population. Response rates to surveys are highly variable and there are indications that, at least in some instances, the rates of response to web-based exchanges are lower than are achieved with paper-based methods [26]. Nevertheless, the costs of administering follow-up at scale are substantially lower using digital technology than traditional print-based methods and the flexibility and potential immediacy of contact between physician and patient have a face-value attraction. Establishing why so many of our patients declined to adopt this option would require in-depth interviewing of several thousand patients and is beyond the resources of the study as originally conceived. Similarly, we are not equipped to investigate whether or how physicians advocated for this aspect of the study during clinic visits or how patients responded to encouragement.

Adherence to therapy for patients self-reporting via the DIAPAsOn digital platform was defined as the total number of days when the patient took the full prescribed dose of OM3EE during the specified period divided by the total number of days in that period. Calculated in that way, adherence appeared to be low in these patients and declined during the period of observation. However, we have no means of ascertaining whether the data that patients recorded accurately reflected their true adherence to study medication: actual adherence rates may therefore have been higher than the recorded findings suggest. Such a possibility would be compatible with the finding that in-study trends in lipid and lipoprotein indices were favorable and numerically very similar in a small group of digital adopters and the rest of the analysis population. We are unable to demonstrate any such alignment of adherence rates, however, finding instead a notable discrepancy between adherence rates registered by users of the DIAPAsOn app and those in whom adherence was assessed only at clinic visits despite broadly similar lipid responses in both subgroups.

Comparison of the digital subset with the main analysis population also identified no demographic differences between the two groups that might explain the adoption/non-adoption of the digital resources of DIAPAsOn. Wide-ranging technical obstacles seem unlikely given the high level of smartphone penetration in Russia [27], general access to the internet and the requirement for digital proficiency as an inclusion condition. Average age (≍58 years) is not, prima facie, a sufficient explanation for the low level of digital uptake but may have exerted an influence that our study was not calibrated to identify.

Seemingly at variance with the low level of adoption of the mobile technology devised for DIAPAsOn—and the apparently low levels of medication adherence reported by those patients that used the technology—is the observation that the dropout rate among the digital adopters was zero. The impression of a subset of patients who are tenacious in their adherence to technology but inattentive to their medications is a paradox that we are at present unable to rationalize.

Another finding of note in the digitally engaged patients was that HRQoL indices showed a striking and sustained improvement among those who self-reported high compliance. Patient numbers were small, the structure of an observational study precludes determination of cause and effect and ascertaining reasons for this improvement lie outside the scope of our research. This is, nevertheless, an intriguing finding that would merit attention in future investigation.

The considered view of mobile- or internet-based health interventions to promote adherence to therapy is that these innovations have potential but that both the quality and range of research need to be enhanced [28–30]. DIAPAsOn should be considered in that context. One lesson that might be extracted from our findings is that patient engagement and participation in the development of an online or mobile telephony-based adherence aid might produce longer-term gains in terms of patients’ willingness to engage with the service once it is brought into daily use. DIAPAsOn might perhaps have benefited from a small-scale scoping study or a pilot phase before the technology was deployed in such a large patient population. Evaluation of our digital platform by means of the Mobile Application Rating Scale [31] might also have helped to refine the technology and enhance its acceptance by patients, and the failure to apply that test in advance may be considered a missed opportunity.

One hazard of such an approach, however, is that by catering to the priorities of patients well-disposed to mobile health technology the needs of ‘digital exiles’ are overlooked. DIAPAsOn required some evidence of individual technical competence from its patients but was otherwise non-restrictive in its eligibility criteria. To that extent it has provided useful insights into the sort of real-world populations that might be encountered (at least in Russia) and some of the challenges that these populations pose for proponents of mobile or ehealth services.

As with observational studies in general, the absence of a control group precludes any determination of cause and effect and the potential for biases in any trial of this type must be acknowledged. A retrospective calculation of the Nichol score [32] for DIAPAsOn confirmed that our study rated favorably in the subcategories ‘Disease-Related Criteria’ and ‘Compliance Definition and Measurement Criteria’ but scored less strongly in the subcategory ‘Study Design Criteria’. The duration of follow-up was appropriate for a first assessment of a technical innovation but a substantially longer period of observation would be needed to demonstrate robust and meaningful improvements in long-term compliance and adherence, regardless of the methods or technologies used.

## Conclusions

Data from DIAPAsOn collected in our study confirm the clinical profile of OM3EE as an effective and well-tolerated lipid-modifying therapy and as an appropriate element of the medical regimen(s) for management of hypertriglyceridemia or the secondary prevention of MI. Substantial (∼1 mmol/L) baseline-dependent reductions in TG were recorded and other nominally advantageous alterations in the lipid profile were apparent, including reduction in levels of non-HDL-C, regardless of the concomitant use of statins and/or fibrates. Investigations into compliance with therapy produced conflicting results depending on the method of reporting used. Uptake of digital methods for self-reporting of adherence to therapy was low and indicates a need for further research into the factors that motivate or discourage patients to take advantage of such services and of how best to use those technologies to promote treatment compliance.

## Data Availability

All relevant data are within the paper and its Supplementary Materials.

## Acknowledgements

The investigators thank the Ethics Committee of Pirogov Russian National Research Medical University for its advice and guidance on the development of the research protocol.

Manuscript preparation was assisted by **Hughes associates**, Oxford, UK.

## Supplementary Tables

**Table S1a.** Assessment of the relationship between the rate of adherence and sex.

| Visit | Parameter | Gender |  |  |  |
| --- | --- | --- | --- | --- | --- |
|  |  | Male |  | Female |  |
|  |  | N | Proportion (%) | N° Pt | Rate (%) |
| Visit 1 –<br>Visit 3 | Total | 1039 | 100 | 936 | 100 |
|  | Low adherence | 1 | 0.1 | 7 | 0.75 |
|  | Moderate adherence | 52 | 5 | 83 | 8.87 |
|  | High adherence | 180 | 17.32 | 214 | 22.86 |
|  | Very high adherence | 806 | 77.57 | 632 | 67.52 |
|  | Mean questionnaire score | 13.23 |  | 12.53 |  |
|  | p-value* | <0.001 |  |  |  |

Note: data are of the PP population, N = 2167; \* $\chi$ -square.

**Table S1b.** Assessment of the relationship between the rate of adherence to therapy and the sex of patients to whom OM3EE was prescribed as secondary preventive therapy after myocardial infarction.

| Visit | Parameter | Gender |  |  |  |
| --- | --- | --- | --- | --- | --- |
|  |  | Male |  | Female |  |
|  |  | N | Proportion (%) | N° Pt | Rate (%) |
| Visit 1 –<br>Visit 3 | Total | 528 | 100 | 252 | 100 |
|  | Low adherence | 0 | 0 | 0 | 0 |
|  | Moderate adherence | 10 | 1.89 | 7 | 2.78 |
|  | High adherence | 58 | 10.98 | 26 | 10.32 |
|  | Very high adherence | 460 | 87.12 | 219 | 86.9 |
|  | Mean questionnaire score | 13.9 |  | 13.9 |  |
|  | p-value* | 0.98 |  |  |  |

Note: data are of the PP population, N = 2167; \* $\chi$ -square.

**Table S1c.** Assessment of the relationship between the rate of adherence to therapy and sex among patients to whom OM3EE was prescribed for hypertriglyceridemia

| Visit | Parameter | Gender |  |  |  |
| --- | --- | --- | --- | --- | --- |
|  |  | Male |  | Female |  |
|  |  | N | Proportion (%) | N | Proportion (%) |
| Visit 1 – Visit 3 | Total | 511 | 100 | 684 | 100 |
|  | Low adherence | 1 | 0.2 | 7 | 1.02 |
|  | Moderate adherence | 42 | 8.22 | 76 | 11.11 |
|  | High adherence | 122 | 23.87 | 188 | 27.49 |
|  | Very high adherence | 346 | 67.71 | 413 | 60.38 |
|  | Mean questionnaire score | 12.54 |  | 12.03 |  |
|  | p-value* | 0.007 |  |  |  |

Note: data are of the PP population, N = 2167; \* $\chi^2$ -square.

**Table S2a.** Assessment of the relationship between the rate of adherence and age.

| Visit | Parameter | Age group |  |  |  |  |  |  |  |
| --- | --- | --- | --- | --- | --- | --- | --- | --- | --- |
|  |  | 18-44 |  | 45-60 |  | 61-75 |  | >75 |  |
|  |  | N | Proportion (%) | N | Proportion (%) | N | Proportion (%) | N | Proportion (%) |
| Visit 1 – Visit 3 | Total | 231 | 100 | 768 | 100 | 804 | 100 | 172 | 100 |
|  | Low adherence | 1 | 0.43 | 3 | 0.39 | 4 | 0.5 | 0 | 0 |
|  | Moderate adherence | 21 | 9.09 | 52 | 6.77 | 48 | 5.97 | 14 | 8.14 |
|  | High adherence | 55 | 23.81 | 167 | 21.74 | 137 | 17.04 | 35 | 20.35 |
|  | Very high adherence | 154 | 66.67 | 546 | 71.09 | 615 | 76.49 | 123 | 71.51 |
|  | Mean questionnaire score | 12.45 |  | 12.78 |  | 13.15 |  | 12.87 |  |
|  | p-value* | 0.007 |  |  |  |  |  |  |  |

Note: data are of the PP population, N = 2167; \* $\chi^2$ -square.

**Table S2b.** Assessment of the relationship between the rate of adherence to therapy and the age of patients to whom OM3EE was prescribed as secondary preventive therapy after myocardial infarction.

| Visit | Parameter | Age group |  |  |  |  |  |  |  |
| --- | --- | --- | --- | --- | --- | --- | --- | --- | --- |
|  |  | 18-44 |  | 45-60 |  | 61-75 |  | >75 |  |
|  |  | N | Proportion (%) | N | Proportion (%) | N | Proportion (%) | N | Proportion (%) |
| Visit 1 – Visit 3 | Total | 48 | 100 | 273 | 100 | 378 | 100 | 81 | 100 |
|  | Low adherence | 0 | 0 | 0 | 0 | 0 | 0 | 0 | 0 |
|  | Moderate adherence | 1 | 2.08 | 3 | 1.1 | 8 | 2.12 | 5 | 6.17 |
|  | High adherence | 5 | 10.42 | 25 | 9.16 | 39 | 10.32 | 15 | 18.52 |
|  | Very high adherence | 42 | 87.5 | 245 | 89.74 | 331 | 87.57 | 61 | 75.31 |
|  | Mean questionnaire score | 13.92 |  | 14.06 |  | 13.95 |  | 13.14 |  |
|  | p-value* | 0.007 |  |  |  |  |  |  |  |

Note: data are of the PP population, N = 2167; \* $\chi$ -square.

**Table S2c.** Assessment of the relationship between the rate of adherence to therapy and age among patients to whom OM3EE was prescribed for hypertriglyceridemia.

| Visit | Parameter | Age group |  |  |  |  |  |  |  |
| --- | --- | --- | --- | --- | --- | --- | --- | --- | --- |
|  |  | 18-44 |  | 45-60 |  | 61-75 |  | >75 |  |
|  |  | N | Proportion (%) | N | Proportion (%) | N | Proportion (%) | N | Proportion (%) |
| Visit 1 – Visit 3 | Total | 183 | 100 | 495 | 100 | 426 | 100 | 91 | 100 |
|  | Low adherence | 1 | 0.55 | 3 | 0.61 | 4 | 0.94 | 0 | 0 |
|  | Moderate adherence | 20 | 10.93 | 49 | 9.9 | 40 | 9.39 | 9 | 9.89 |
|  | High adherence | 50 | 27.32 | 142 | 28.69 | 98 | 23 | 20 | 21.98 |
|  | Very high adherence | 112 | 61.2 | 301 | 60.81 | 284 | 66.67 | 62 | 68.13 |
|  | Mean questionnaire score | 12.07 |  | 12.07 |  | 12.44 |  | 12.64 |  |
|  | p-value* | 0.19 |  |  |  |  |  |  |  |

Note: data are of the PP population, N = 2167; \* $\chi$ -square.

**Table S3a.** Assessment of the relationship between the rate of adherence and work status.

| Visit | Parameter | Work status |  |  |  |
| --- | --- | --- | --- | --- | --- |
|  |  | Works |  | Does not work |  |
|  |  | N | Proportion (%) | N | Proportion (%) |
| Visit 1 – Visit 3 | Total | 1096 | 100 | 879 | 100 |
|  | Low adherence | 5 | 0.46 | 3 | 0.34 |
|  | Moderate adherence | 85 | 7.76 | 50 | 5.69 |
|  | High adherence | 251 | 22.9 | 143 | 16.27 |
|  | Very high adherence | 755 | 68.89 | 683 | 77.7 |
|  | Mean questionnaire score | 12.66 |  | 13.2 |  |
|  | p-value* | <0.001 |  |  |  |

Note: data are of the PP population, N = 2167; \* $\chi$ -square.

**Table S3b.** Assessment of the relationship between the rate of adherence to therapy and work status of patients to whom OM3EE was prescribed as secondary preventive therapy after myocardial infarction.

| Visit | Parameter | Work status |  |  |  |
| --- | --- | --- | --- | --- | --- |
|  |  | Works |  | Does not work |  |
|  |  | N | Proportion (%) | N | Proportion (%) |
| Visit 1 – Visit 3 | Total | 382 | 100 | 398 | 100 |
|  | Low adherence | 0 | 0 | 0 | 0 |
|  | Moderate adherence | 6 | 1.57 | 11 | 2.76 |
|  | High adherence | 47 | 12.3 | 37 | 9.3 |
|  | Very high adherence | 329 | 86.13 | 350 | 87.94 |
|  | Mean questionnaire score | 13.9 |  | 13.9 |  |
|  | p-value * | 0.99 |  |  |  |

Note: data are of the PP population, N = 2167; \* $\chi$ -square.

**Table S3c.** Assessment of the relationship between the rate of adherence to therapy and work status among patients to whom OM3EE was prescribed for hypertriglyceridemia.

| Visit | Parameter | Work status |  |  |  |
| --- | --- | --- | --- | --- | --- |
|  |  | Works |  | Does not work |  |
|  |  | N | Proportion (%) | N | Proportion (%) |
| Visit 1 – Visit 3 | Total | 714 | 100 | 481 | 100 |
|  | Low adherence | 5 | 0.7 | 3 | 0.62 |
|  | Moderate adherence | 79 | 11.06 | 39 | 8.11 |
|  | High adherence | 204 | 28.57 | 106 | 22.04 |
|  | Very high adherence | 426 | 59.66 | 333 | 69.23 |
|  | Mean questionnaire score | 11.99 |  | 12.62 |  |
|  | p-value* | 0.001 |  |  |  |

Note: data are of the PP population, N = 2167; \* $\chi$ -square.

**Table S4a.** Assessment of the relationship between the rate of adherence and education.

| Visit | Parameter | Education |  |  |  |  |  |  |  |  |  |
| --- | --- | --- | --- | --- | --- | --- | --- | --- | --- | --- | --- |
|  |  | Secondary education drop-out |  | Secondary general |  | Secondary vocational |  | Higher |  | Supplementary vocational |  |
|  |  | N | Proportion (%) | N | Proportion (%) | N | Proportion (%) | N | Proportion (%) | N | Proportion (%) |
| Visit 1 – Visit 3 | Total | 15 | 100 | 224 | 100 | 634 | 100 | 1064 | 100 | 38 | 100 |
|  | Low adherence | 0 | 0 | 3 | 1.34 | 3 | 0.47 | 2 | 0.19 | 0 | 0.19 |
|  | Moderate adherence | 0 | 0 | 24 | 10.71 | 56 | 8.83 | 52 | 4.89 | 3 | 4.89 |
|  | High adherence | 1 | 6.67 | 67 | 29.91 | 121 | 19.09 | 192 | 18.05 | 13 | 18.05 |
|  | Very high adherence | 14 | 93.33 | 130 | 58.04 | 454 | 71.61 | 818 | 76.88 | 22 | 76.88 |
|  | Mean questionnaire score | 14.27 |  | 11.91 |  | 12.67 |  | 13.25 |  | 12.21 |  |
|  | p-value* | <0.001 |  |  |  |  |  |  |  |  |  |

Note: data are of the PP population, N = 2167; \* $\chi$ -square.

**Table S4b.** Assessment of the relationship between the rate of adherence to therapy and education of patients to whom OM3EE was prescribed as secondary preventive therapy after myocardial nfarction.

| Visit | Parameter | Education |  |  |  |  |  |  |  |  |  |
| --- | --- | --- | --- | --- | --- | --- | --- | --- | --- | --- | --- |
|  |  | Secondary education drop-out |  | Secondary general |  | Secondary vocational |  | Higher |  | Supplementary vocational |  |
|  |  | N | Proportion (%) | N | Proportion (%) | N | Proportion (%) | N | Proportion (%) | N | Proportion (%) |
| Visit 1 – Visit 3 | Total | 12 | 100 | 78 | 100 | 258 | 100 | 419 | 100 | 13 | 100 |
|  | Low adherence | 0 | 0 | 0 | 0 | 0 | 0 | 0 | 0 | 0 | 0 |
|  | Moderate adherence | 0 | 0 | 3 | 3.85 | 8 | 3.1 | 6 | 1.43 | 0 | 1.43 |
|  | High adherence | 1 | 8.33 | 12 | 15.38 | 24 | 9.3 | 46 | 10.98 | 1 | 10.98 |
|  | Very high adherence | 11 | 91.67 | 63 | 80.77 | 226 | 87.6 | 367 | 87.59 | 12 | 87.59 |
|  | Mean questionnaire score | 14.17 |  | 13.46 |  | 13.84 |  | 14.01 |  | 14.15 |  |
|  | p-value* | 0.3 |  |  |  |  |  |  |  |  |  |

Note: data are of the PP population, N = 2167; \* $\chi$ -square.

**Table S4c.** Assessment of the relationship between the rate of adherence to therapy and education among patients to whom OM3EE was prescribed for hypertriglyceridemia.

| Visit | Parameter | Education |  |  |  |  |  |  |  |  |  |
| --- | --- | --- | --- | --- | --- | --- | --- | --- | --- | --- | --- |
|  |  | Secondary education drop-out |  | Secondary general |  | Secondary vocational |  | Higher |  | Supplementary vocational |  |
|  |  | N | Proportion (%) | N | Proportion (%) | N | Proportion (%) | N | Proportion (%) | N | Proportion (%) |
| Visit 1 – Visit 3 | Total | 3 | 100 | 146 | 100 | 376 | 100 | 645 | 100 | 25 | 100 |
|  | Low adherence | 0 | 0 | 3 | 2.05 | 3 | 0.8 | 2 | 0.31 | 0 | 0.31 |
|  | Moderate adherence | 0 | 0 | 21 | 14.38 | 48 | 12.77 | 46 | 7.13 | 3 | 7.13 |
|  | High adherence | 0 | 0 | 55 | 37.67 | 97 | 25.8 | 146 | 22.64 | 12 | 22.64 |
|  | Very high adherence | 3 | 100 | 67 | 45.89 | 228 | 60.64 | 451 | 69.92 | 10 | 69.92 |
|  | Mean questionnaire score | 14.67 |  | 11.08 |  | 11.88 |  | 12.76 |  | 11.2 |  |
|  | p-value* | <0.001 |  |  |  |  |  |  |  |  |  |

Note: data are of the PP population, N = 2167; \* $\chi$ -square.

**Table S5a.** Assessment of the relationship between the rate of adherence and marital status.

| Visit | Parameter | Marital status |  |  |  |  |  |  |  |
| --- | --- | --- | --- | --- | --- | --- | --- | --- | --- |
|  |  | Single |  | Married |  | Divorced |  | Widowed |  |
|  |  | N | Proportion (%) | N | Proportion (%) | N | Proportion (%) | N | Proportion (%) |
| Visit 1 –<br>Visit 3 | Total | 82 | 100 | 1593 | 100 | 114 | 100 | 186 | 100 |
|  | Low adherence | 0 | 0 | 7 | 0.44 | 1 | 0.88 | 0 | 0 |
|  | Moderate adherence | 13 | 15.85 | 102 | 6.4 | 8 | 7.02 | 12 | 6.45 |
|  | High adherence | 13 | 15.85 | 320 | 20.09 | 24 | 21.05 | 37 | 19.89 |
|  | Very high adherence | 56 | 68.29 | 1164 | 73.07 | 81 | 71.05 | 137 | 73.66 |
|  | Mean questionnaire score | 12.5 |  | 12.91 |  | 12.76 |  | 13.08 |  |
|  | p-value* | 0.49 |  |  |  |  |  |  |  |

Note: data are of the PP population, N = 2167; \* $\chi$ -square.

**Table S5b.** Assessment of the relationship between the rate of adherence to therapy and marital status of patients to whom OM3EE was prescribed as secondary preventive therapy after myocardial infarction.

| Visit | Parameter | Marital status |  |  |  |  |  |  |  |
| --- | --- | --- | --- | --- | --- | --- | --- | --- | --- |
|  |  | Single |  | Married |  | Divorced |  | Widowed |  |
|  |  | N | Proportion (%) | N | Proportion (%) | N | Proportion (%) | N | Proportion (%) |
| Visit 1 – Visit 3 | Total | 31 | 100 | 574 | 100 | 57 | 100 | 118 | 100 |
|  | Low adherence | 0 | 0 | 0 | 0 | 0 | 0 | 0 | 0 |
|  | Moderate adherence | 4 | 12.9 | 7 | 1.22 | 3 | 5.26 | 3 | 2.54 |
|  | High adherence | 6 | 19.35 | 55 | 9.58 | 5 | 8.77 | 18 | 15.25 |
|  | Very high adherence | 21 | 67.74 | 512 | 89.2 | 49 | 85.96 | 97 | 82.2 |
|  | Mean questionnaire score | 12.58 |  | 14.01 |  | 13.79 |  | 13.8 |  |
|  | p-value* | 0.004 |  |  |  |  |  |  |  |

Note: data are of the PP population, N = 2167; \* $\chi$ -square.

**Table S5c.**
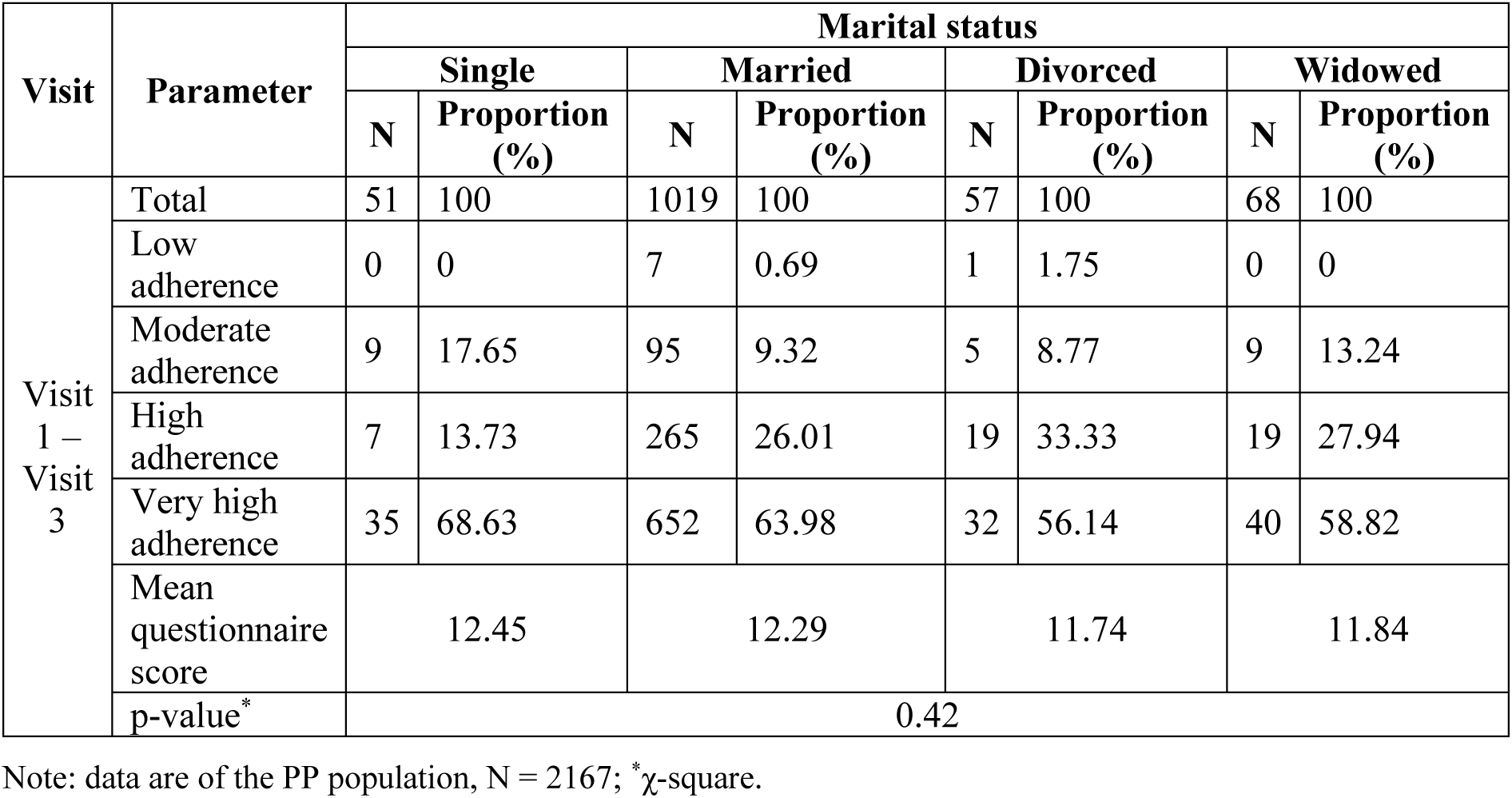
Assessment of the relationship between the rate of adherence to therapy and marital status among patients to whom OM3EE was prescribed for hypertriglyceridemia.

| Visit | Parameter | Marital status |  |  |  |  |  |  |  |
| --- | --- | --- | --- | --- | --- | --- | --- | --- | --- |
|  |  | Single |  | Married |  | Divorced |  | Widowed |  |
|  |  | N | Proportion (%) | N | Proportion (%) | N | Proportion (%) | N | Proportion (%) |
| Visit 1 – Visit 3 | Total | 51 | 100 | 1019 | 100 | 57 | 100 | 68 | 100 |
|  | Low adherence | 0 | 0 | 7 | 0.69 | 1 | 1.75 | 0 | 0 |
|  | Moderate adherence | 9 | 17.65 | 95 | 9.32 | 5 | 8.77 | 9 | 13.24 |
|  | High adherence | 7 | 13.73 | 265 | 26.01 | 19 | 33.33 | 19 | 27.94 |
|  | Very high adherence | 35 | 68.63 | 652 | 63.98 | 32 | 56.14 | 40 | 58.82 |
|  | Mean questionnaire score | 12.45 |  | 12.29 |  | 11.74 |  | 11.84 |  |
|  | p-value* | 0.42 |  |  |  |  |  |  |  |

Note: data are of the PP population, N = 2167; \* $\chi^2$ -square.

## Appendix 1. Listing of DIAPASoN center investigators.

| Region | Investigator |
| --- | --- |
| Astrakhan Oblast | 1. Borodulya Mihail Vasil'evich |
| Bryansk Oblast | 2. Petuhova Irina Leonidovna |
| Vladimir Oblast | 3. Timofeeva Irina Vladimirovna |
| Volgograd Oblast | 4. Vorob'eva Svetlana Gennad'evna |
|  | 5. Gulova Ol'ga Aleksandrovna |
|  | 6. Dugencova Larisa Anatol'evna |
|  | 7. Sytilina Natal'ya Nikolaevna |
| Voronezh Oblast | 8. Trubicyna Irina Valentinovna |
| Kemerovo Oblast | 9. Krestova Ol'ga Sergeevna |
| Krasnodar Krai | 10. Alekseeva Elena Valer'evna |
|  | 11. Allaabed Hassan Adnanovich |
|  | 12. Bahmet'eva Irina Aleksandrovna |
|  | 13. Boldin Vasilij Borisovich |
|  | 14. Vikulova Larisa Vladimirovna |
|  | 15. Golovko Artyom Yur'evich |
|  | 16. Emuzova Liana Anatol'evna |
|  | 17. Zaharov Aleksandr Yur'evich |
|  | 18. Ivochkina Marina Ivanovna |
|  | 19. Imamutdinova Marina Vladimirovna |
|  | 20. Kovalenko Fyodor Andreevich |
|  | 21. Minasyan Ani Kamoevna |
|  | 22. Mkrtycheva Angelina Gennad'evna |
|  | 23. Nikitina Elena Valer'evna |
|  | 24. Novikova Svetlana Valer'evna |
|  | 25. Pavlovec Vadim Petrovich |
|  | 26. Perkova Elena Mihajlovna |
|  | 27. Prozorovskaya Yuliya Igorevna |
|  | 28. Ramenskaya Tat'yana Evgen'evna |
|  | 29. Raff Stanislav Anatol'evich |
|  | 30. Smolina Elena Garievna |
|  | 31. Subbotina Anastasiya Vladimirovna |
|  | 32. Tatarinceva Zoya Gennad'evna |
|  | 33. Usmanova Natal'ya Aleksandrovna |
|  | 34. Hazhbieva Milana Musaevna |
|  | 35. Shadzhe Evgeniya Azamatovna |
| Krasnoyarsk Krai | 36. Hamyt- Kyzy Ajperi Hamytovna |
| Moscow | 37. Adamyan Margarita Mamikonovna |
|  | 38. Belov Leonid L'vovich |
|  | 39. Brodichko Alena Ivanovna |
|  | 40. Bugaev Timofej Dmitrievich |
|  | 41. Vahrulina Natal'ya Konstantinovna |
|  | 42. Grigor'eva Ekaterina Anatol'evna |
|  | 43. Dvorina Ol'ga Gennad'evna |
|  | 44. Dmitrieva Irina Mihajlovna |
|  | 45. Karaeva Aida Anzorovna |
|  | 46. Novosel'ceva Ekaterina Pavlovna |
|  | 47. Oganesyan Lala Konstantinovna |
|  | 48. Polyakova Natal'ya Olegovna |
|  | 49. Ryzhova Tat'yana Vladimirovna |
|  | 50. Smirnova Ol'ga L'vovna |
|  | 51. Tavleeva Svetlana Nikolaevna |
|  | 52. Hrulenko Svetlana Borisovna |
|  | 53. Chernushenko Tat'yana Ivanovna |
| Moscow Oblast | 54. Gukov Konstantin Aleksandrovich |
|  | 55. Gukova Tat'yana Sergeevna |
|  | 56. Malyarenko Elena Nikolaevna |
|  | 57. Sorokin Sergej Anatol'evich |
| Nizhny Novgorod Oblast | 58. Abramova Nataliya Arkad'evna |
|  | 59. Aksenova Nataliya Aleksandrovna |
|  | 60. Barsukova Nataliya Aleksandrovna |
|  | 61. Budarina Yuliya Gennad'evna |
|  | 62. Grushin Dmitrij Valer'evich |
|  | 63. Kolesnichenko Irina Vyacheslavovna |
|  | 64. Lokonova Larisa Mihajlovna |
|  | 65. Fedorova Svetlana Nikolaevna |
| Novosibirsk Oblast | 66. Shurkevich Anastasiya Alekseevna |
| Omsk Oblast | 67. Minzhasarova Saniya Hasangalievna |
|  | 68. Naumov Dmitrij Valer'evich |
| Perm Krai | 69. Zhuravleva Natal'ya Alekseevna |
| The Republic of Adygea | 70. Shekhmirzova Dzhanetta Ruslanovna |
| The Republic of Bashkortostan | 71. Gilyaeva El'vira Fanisovna |
|  | 72. Dmitriev Aleksej Valer'evich |
|  | 73. Murasova Rimma Ismagilovna |
|  | 74. Tarzimanova Yuliya Shamilevna |
|  | 75. Timerbulatov Timur Rasfarovich |
| Tatarstan | 76. Ivanova Natal'ya Mihajlovna |
| Rostov Oblast | 77. Budanova Ol'ga Veniaminovna |
|  | 78. Lobe Aleksandra Ovanesovna |
|  | 79. Mazruho Marina Karpovna |
|  | 80. Morgun Nina Karpovna |
|  | 81. Stupina Anna Alekseevna |
| Ryazan Oblast | 82. Grusheckaya Irina Stanislavovna |
|  | 83. Samarceva Yana Nikolaevna |
| Samara Oblast | 84. Aristova Tat'yana Vladimirovna |
|  | 85. Reznik Irina Mihajlovna |
|  | 86. Rybina Evgeniya Dmitrievna |
|  | 87. Sapunkova Svetlana Mihajlovna |
|  | 88. Filippovskaya Natal'ya Igorevna |
|  | 89. Chernova Viktoriya Nikolaevna |
| Saint Petersburg | 90. Bulycheva-Samohina Lyubov' Vasil'evna |
|  | 91. Omel'chenko Marina Yur'evna |
|  | 92. Saf'yanova Natal'ya Viktorovna |
| Saratov Oblast | 93. Mihajlova Elena Aleksandrovna |
| Smolensk Oblast | 94. Novik Lyudmila Mihajlovna |
| Stavropol Krai | 95. Vedeneva Elena Viktorovna |
|  | 96. Eremenko Aleksej Mihajlovich |
|  | 97. Kubanova Asiyat Borisovna |
|  | 98. Minasova Elena Nikolaevna |
| Tomsk Oblast | 99. Zubova Ol'ga Valer'evna |
| Tula Oblast | 100. Barabanova Tat'yana Yur'evna |
|  | 101. Dabizha Viktoriya Gennad'evna |
|  | 102. Kolomejceva Tat'yana Mihajlovna |
|  | 103. Prihod'ko Tat'yana Nikolaevna |
| Tumen Oblast | 104. Ahshiyatova Nastya Ibragimovna |
| Khabarovsk Krai | 105. Koroleva Ramilya Lotfullovna |
| Chelyabinsk Oblast | 106. Malyutina Anastasiya Gennad'evna |
|  | 107. Fanina El'vira Rinatovna |

## References

1. Kones R. Molecular sources of residual cardiovascular risk, clinical signals, and innovative solutions: relationship with subclinical disease, undertreatment, and poor adherence: implications of new evidence upon optimizing cardiovascular patient outcomes. Vasc Health Risk Manag. 2013;9:617–670. doi:10.2147/VHRM.S37119

2. Schwartz GG, Abt M, Bao W, DeMicco D, Kallend D, Miller M, et al. Fasting triglycerides predict recurrent ischemic events in patients with acute coronary syndrome treated with statins. J Am Coll Cardiol 2015;65:2267e75.

3. Hussain A, Ballantyne CM, Saeed A, Virani SS. Triglycerides and ASCVD Risk Reduction: Recent Insights and Future Directions. Curr Atheroscler Rep. 2020;22(7):25. Published 2020 Jun 3. doi:10.1007/s11883-020-00846-8

4. Morrison A, Hokanson JE. The independent relationship between triglycerides and coronary heart disease. Vasc Health Risk Manag 2009:5 89–95.

5. Emerging Risk Factors Collaboration, Di Angelantonio E, Sarwar N, Perry P, Kaptoge S, Ray KK, Thompson A, Wood AM, Lewington S, Sattar N, Packard CJ, Collins R, Thompson SG, Danesh J. Major lipids, apolipoproteins,and risk of vascular disease. JAMA 2009;302:1993–2000.

6. Lorenzatti AJ, Toth PP. New Perspectives on Atherogenic Dyslipidaemia and Cardiovascular Disease. Eur Cardiol. 2020;15:1–9. Published 2020 Feb 26. doi:10.15420/ecr.2019.06

7. Vallejo-Vaz AJ, Corral P, Schreier L, Ray KK. Triglycerides and residual risk. Curr Opin Endocrinol Diabetes Obes. 2020;27(2):95–103. doi:10.1097/MED.0000000000000530

8. Miller M, Stone NJ, Ballantyne C, et al. Triglycerides and cardiovascular disease: a scientific statement from the American Heart Association. Circulation. 2011;123(20):2292–2333. doi:10.1161/CIR.0b013e3182160726

9. Fan W, Philip S, Granowitz C, Toth PP, Wong ND. Residual Hypertriglyceridemia and Estimated Atherosclerotic Cardiovascular Disease Risk by Statin Use in U.S. Adults With Diabetes: National Health and Nutrition Examination Survey 2007-2014. Diabetes Care. 2019;42(12):2307–2314. doi:10.2337/dc19-0501

10. Brown WV, Jacobson TA, Braun LT. Achieving adherence to lipid-lowering regimens. J Clin Lipidol. 2013;7(1):4–13. doi:10.1016/j.jacl.2012.11.004

11. Tsuyuki RT, Bungard TJ. Poor adherence with hypolipidemic drugs: a lost opportunity. Pharmacotherapy. 2001;21(5):576–582. doi:10.1592/phco.21.6.576.34541

12. lakunchykova O, Averina M, Wilsgaard T, et al. Why does Russia have such high cardiovascular mortality rates? Comparisons of blood-based biomarkers with Norway implicate non-ischaemic cardiac damage. J Epidemiol Community Health Published Online First: 15 May 2020. doi: 10.1136/jech-2020-213885.

13. Boytsov S, Logunova N, Khomitskaya Y; CEPHEUS II investigators. Suboptimal control of lipid levels: results from the non-interventional Centralized Pan-Russian Survey of the Undertreatment of Hypercholesterolemia II (CEPHEUS II). Cardiovasc Diabetol. 2017;16(1):158. Published 2017 Dec 16. doi:10.1186/s12933-017-0641

14. Cybulsky M, Cook S, Kontsevaya AV, Vasiljev M, Leon DA. Pharmacological treatment of hypertension and hyperlipidemia in Izhevsk, Russia. BMC Cardiovasc Disord. 2016;16:122. Published 2016 Jun 3. doi:10.1186/s12872-016-0300-9

15. Toth PP, Granowitz C, Hull M, Anderson A, Philip S. Long-term statin persistence is poor among high-risk patients with dyslipidemia: a real-world administrative claims analysis. Lipids Health Dis. 2019;18(1):175. Published 2019 Sep 16. doi:10.1186/s12944-019-1099-z

16. Huber CA, Meyer MR, Steffel J, Blozik E, Reich O, Rosemann T. Post-myocardial Infarction (MI) Care: Medication Adherence for Secondary Prevention After MI in a Large Real-world Population. Clin Ther. 2019;41(1):107–117. doi:10.1016/j.clinthera.2018.11.012

17. Gutstein AS, Copple T. Cardiovascular disease and omega-3s: Prescription products and fish oil dietary supplements are not the same. J Am Assoc Nurse Pract. 2017;29(12):791–801. doi:10.1002/2327-6924.12535

18. Abdelhamid AS, Brown TJ, Brainard JS, et al. Omega-3 fatty acids for the primary and secondary prevention of cardiovascular disease. Cochrane Database Syst Rev. 2020;3(2):CD003177. Published 2020 Feb 29. doi:10.1002/14651858.CD003177.pub5

19. Redfern J. Smart health and innovation: facilitating health-related behaviour change. Proc Nutr Soc. 2017;76(3):328–332. doi:10.1017/S0029665117001094

20. Arutyunov GP, Arutyunov AG, Ageev FT, Fofanova TV. Use of digital technology tools to examine patient adherence to a prescription omega-3 polyunsaturated acids therapy intended to mitigate cardiovascular risk: protocol and preliminary demographic data from the DIAPAsOn prospective observational study. J Med Internet Res Prot. JMIR Res Protoc. 2021 Aug 30;10(8):e29061. doi: 10.2196/29061. PMID: 34459746; PMCID: PMC8438613

21. Fofanova T.V., Ageev F.T., Smirnova M.D., Svirida O.N., Kuzmina A.E., Thostov A.Sh., Nelyubina A.S. National questionnaire of treatment compliance: testing and application in outpatient practice // Systemic hypertension | #2 | 2014, p. 13–16

22. Gaisenok O, Martsevich S, Tripkosh S, Lukina Y. Analysis of lipid-lowering therapy and factors affecting regularity of statin intake in patients with cardiovascular disease enrolled in the PROFILE registry. Rev Port Cardiol. 2015;34(2):111–116. doi:10.1016/j.repc.2014.08.021

23. Ivers NM, Schwalm JD, Bouck Z, et al. Interventions supporting long term adherence and decreasing cardiovascular events after myocardial infarction (ISLAND): pragmatic randomised controlled trial. BMJ. 2020;369:m1731. Published 2020 Jun 10. doi:10.1136/bmj.m1731

24. Drozdova LY, Martsevich SY, Voronina VP. Evaluation of cardiovascular risk factors prevalence and efficacy of their correction in physicians. Estimation of physicians’ expertise in up-to-date clinical guidelines. Results of the “Physician’s health and education” study. Ration Pharmacother Cardiol. 2011;7(2):137–144 [Russian]

25. Bradley CK, Wang TY, Li S, et al. Patient-Reported Reasons for Declining or Discontinuing Statin Therapy: Insights From the PALM Registry. J Am Heart Assoc. 2019;8(7):e011765. doi:10.1161/JAHA.118.011765

26. Sebo P, Maisonneuve H, Cerutti B, Fournier JP, Senn N, Haller DM. Rates, Delays, and Completeness of General Practitioners’ Responses to a Postal Versus Web-Based Survey: A Randomized Trial. J Med Internet Res. 2017;19(3):e83. Published 2017 Mar 22. doi:10.2196/jmir.6308

27. https://www.statista.com/statistics/467166/forecast-of-smartphone-users-in-russia/

28. Gandapur Y, Kianoush S, Kelli HM, et al. The role of mHealth for improving medication adherence in patients with cardiovascular disease: a systematic review. Eur Heart J Qual Care Clin Outcomes. 2016;2(4):237–244. doi:10.1093/ehjqcco/qcw018

29. Hamine S, Gerth-Guyette E, Faulx D, Green BB, Ginsburg AS. Impact of mHealth chronic disease management on treatment adherence and patient outcomes: a systematic review. J Med Internet Res. 2015;17(2):e52. Published 2015 Feb 24. doi:10.2196/jmir.3951

30. Adler AJ, Martin N, Mariani J, et al. Mobile phone text messaging to improve medication adherence in secondary prevention of cardiovascular disease. Cochrane Database Syst Rev. 2017;4(4):CD011851. Published 2017 Apr 29. doi:10.1002/14651858.CD011851.pub2

31. Stoyanov SR, Hides L, Kavanagh DJ, Zelenko O, Tjondronegoro D, Mani M. Mobile app rating scale: a new tool for assessing the quality of health mobile apps. JMIR Mhealth Uhealth. 2015;3(1):e27. doi: 10.2196/mhealth.3422.

32. Nichol MB, Venturini F, Sung JCY. A critical evaluation of the methodology of the literature on medication compliance. Ann Pharmacother 1999;33:531–40.

